# Variability in Coronavirus Disease-2019 Case, Death, and Testing Rates in the United States and Worldwide

**DOI:** 10.1101/2020.10.13.20172957

**Authors:** Ernst J. Schaefer, Andrew S. Geller, Latha Dulipsingh, Margaret R. Diffenderfer, Jeffrey Wisotzkey, Steven B. Kleiboeker

**Affiliations:** Boston Heart Diagnostics/Eurofins Scientific Network, Framingham, MA 01702, USA; Cardiovascular Nutrition Laboratory, Human Nutrition Research Center on Aging at Tufts University, and Tufts University School of Medicine, Boston, MA 02111, USA; Saint Francis Hospital, Trinity Health of New England, Hartford, CT 06105, USA; University of Connecticut School of Medicine, Farmington, CT 06032, USA; Diatherix Laboratories/Eurofins Scientific Network, Huntsville, AL 35806, USA; Viracor-Eurofins Clinical Diagnostics and Scientific Network, Lee’s Summit, MO 64086, USA

**Keywords:** SARS-CoV-2, COVID-19, swab RNA testing

## Abstract

Severe acute respiratory syndrome coronavirus-2 (SARS-CoV-2) infection has been associated with a worldwide pandemic. We assessed data as of November 25, 2020 from our combined laboratories and as reported for states in the United States (US) and countries for case, death, and testing rates per million to determine causes of rate differences. SARS-CoV-2 naso-pharyngeal (NP) RNA testing in 1,179,912 subjects in 47 states (39 of which with >100 cases) are reported, with a mean 9.3% positive rate, comparable to the 7.0% rate reported nationwide. In 91 previously positive (2-4 weeks) subjects, NP swab testing was twice as likely to be positive (58.6%) as saliva samples (21.5%). We also documented that NP swabs could remain positive for 6 weeks or longer. Our positive rates per state agreed reasonably well with reported national data (r=0.609, *P*<0.0001). The highest US case rates per million were in the mid-west; the highest death and testing rates were in the northeast. Of 47 countries, the highest case, death, and testing rates per million were mainly in Europe and the Americas, with the lowest rates in Asia. Correlations between case and death rates and case and testing rates were very different between states (0.076 and -0.093, respectively) and countries (0.763 and 0.600, respectively). In conclusion, outpatient saliva testing was not as sensitive as NP testing for detection, and the marked variability in case and death rates was most likely due to differences in public health measures, viral and human genetic differences and age of cases, rather than due to differences in testing rates.

## INTRODUCTION

Coronavirus disease-2019 (COVID-19) is caused by the severe acute respiratory syndrome coronavirus-2 (SARS-CoV-2) associated with a worldwide pandemic. The diagnosis is made by SARS-CoV-2 RNA detection in naso-pharyngeal (NP) swabs, nasal swabs, oro-pharyngeal (OP) swabs, or saliva.^1-4^ The greatest number of deaths/million in the population has been reported in the northeastern United States. Up to 50% of SARS-CoV-2 positive patients can remain symptomatic; however, such individuals can spread infections.^5-7^ The average onset of symptoms after infection is about 5 days (range 2-14 days). COVID-19 fatality is substantially higher in the elderly and in those with cardiovascular disease, diabetes, obesity, hypertension, and lung disease.

COVID-19 disease symptoms include fever, fatigue, cough, loss of smell and taste, gastro-intestinal symptoms, and shortness of breath. The virus spreads between people mainly via respiratory droplets. Complications include severe acute respiratory syndrome and potentially death from overwhelming infection and inflammation.^1-4^ While testing is critical for diagnosis, public health measures (e.g. face masks, social distancing and quarantining, hand washing, and contact tracing) are critical for prevention of new cases even after widespread vaccinations becomes available. Subjects that are positive for SARS-CoV-2 RNA based on NP swabs may not have transmissible virus over time, but may have only viral fragments in their nasal passages.^7^ Our goals were to assess data obtained from over a million SARS-CoV-2 RNA tests performed in our respective laboratories, as well as available US and worldwide data, in terms of cases, deaths, and testing per million in the population, in order to examine potential causes for the large rate differences observed between states and countries.

## METHODS

### Populations studied

A total of 1,179,912 subjects (58.2% female; age range 1-101 years; median [IQR] age 49.0 [35.0-6.0] years; 18.2% ≥65 years of age) were assessed in physician offices, clinics, and hospitals. These subjects had NP, OP, or nasal swab samples collected by healthcare providers at various sites throughout the United States, placed in viral transport media, and submitted by overnight express courier service for SARS-CoV-2 RNA detection to Boston Heart Diagnostics (Framingham, MA) beginning on April 17, 2020, Diatherix (Huntsville, AL) beginning on March 16, 2020, and/or Viracor (Lee’s Summit, MO) beginning on March 13, 2020. For this analysis, data assessment was ended as of November 1, 2020. Table 1 presents data from hospitals, clinic sites, and healthcare provider offices in 39 states with more than 100 results for samples sent to these laboratories. For this research, patient data were extracted from medical records without name or identification number and were analyzed as anonymized data, determined exempt from institutional review board approval by Advarra Institutional Review Board (Columbia, MD). In our view, this research is exempted from requirement for human institutional review board approval as per exemption 4, as listed at https://grants.nih.gov/policy/humansubjects.htm and at the open education resource (OER) website for research involving human subjects. This exemption “involves the collection or study of data or specimens if publicly available or recorded such that subjects cannot be identified”. We had this designation and our research reviewed by the Advarra Institutional Review Board (Columbia, MD) and their determination was that “had the request for exempt determination been submitted prior to initiation of research activities, the research would have met the criteria for exemption from institutional review board review under 45 CFR 46.104(d). Therefore, they agreed that this research did not require institutional review board approval.

**TABLE 1.** SARS-CoV-2 RNA positive rates by states with >100 tests (Eurofins testing)

| State | Centers for Disease Control and Prevention* |  |  |  | US Eurofins Laboratories<sup>†</sup> |  |
| --- | --- | --- | --- | --- | --- | --- |
|  | Cases/1 M | Deaths/1M | Tests/1M | % Positive<sup>‡</sup> | Tests Done | % Positive<sup>‡</sup> |
| Alabama | 49,347 | 729 | 333,210 | 14.8% | 89,040 | 11.0% |
| Arizona | 43,184 | 902 | 348,958 | 12.4% | 954 | 8.6% |
| Arkansas | 50,575 | 807 | 589,831 | 8.6% | 17,529 | 7.9% |
| California | 29,718 | 482 | 584,547 | 5.1% | 52,275 | 2.9% |
| Colorado | 37,627 | 513 | 293,408 | 8.8% | 8,723 | 6.5% |
| Connecticut | 30,615 | 1,382 | 862,624 | 3.5% | 565 | 3.5% |
| Delaware | 34,484 | 782 | 422,132 | 8.2% | 5,784 | 4.9% |
| Florida | 44,775 | 850 | 566,357 | 8.0% | 124,733 | 16.1% |
| Georgia | 43,468 | 879 | 431,979 | 10.0% | 181,136 | 13.8% |
| Idaho | 54,001 | 501 | 390,728 | 13.8% | — | — |
| Illinois | 55,043 | 994 | 805,890 | 6.8% | 11,891 | 10.4% |
| Indiana | 47,368 | 826 | 609,049 | 7.8% | 11,316 | 8.3% |
| Iowa | 70,615 | 733 | 378,983 | 18.6% | 2,615 | 7.5% |
| Kansas | 51,470 | 516 | 274,322 | 18.8% | 23,043 | 7.3% |
| Kentucky | 40,075 | 427 | 624,592 | 6.4% | 43,881 | 9.0% |
| Louisiana | 48,537 | 1,366 | 730,872 | 6.6% | 12,823 | 9.1% |
| Maryland | 31,507 | 752 | 709,287 | 4.4% | 1,303 | 7.4% |
| Massachusetts | 31,134 | 1,538 | 1,179,643 | 2.6% | 2,838 | 8.0% |
| Michigan | 35,290 | 918 | 683,096 | 5.2% | 29,680 | 10.2% |
| Minnesota | 51,298 | 609 | 701,632 | 7.3% | — | — |
| Mississippi | 47,004 | 1,264 | 432,712 | 10.9% | 4,271 | 20.2% |
| Missouri | 49,400 | 657 | 518,632 | 9.5% | 71,684 | 3.5% |
| Montana | 55,841 | 616 | 591,261 | 9.4% | — | — |
| Nebraska | 62,074 | 506 | 373,945 | 16.5% | 11,4477 | 12.0% |
| Nevada | 47,004 | 680 | 517,154 | 9.1% | — | — |
| New Jersey | 37,042 | 1,920 | 656,070 | 5.6% | 2,846 | 7.7% |
| New Mexico | 42,825 | 701 | 717,682 | 6.0% | — | — |
| New York | 33,835 | 1,768 | 963,495 | 3.5% | 213,926 | 4.3% |

| State | Centers for Disease Control and Prevention* |  |  |  | US Eurofins Laboratories† |  |
| --- | --- | --- | --- | --- | --- | --- |
|  | Cases/1 M | Deaths/1M | Tests/1M | % Positive‡ | Tests Done | % Positive‡ |
| North Carolina | 33,038 | 490 | 482,971 | 6.8% | 74,880 | 7.2% |
| North Dakota | 100,309 | 1,177 | 452,396 | 22.2% | – | – |
| Ohio | 32,774 | 537 | 499,249 | 6.6% | 21,959 | 10.7% |
| Oklahoma | 46,587 | 425 | 523,617 | 7.5% | 9,937 | 13.0% |
| Oregon | 16,598 | 209 | 244,364 | 6.8% | 2,109 | 6.9% |
| Pennsylvania | 26,636 | 804 | 271,595 | 9.8% | 11,377 | 4.1% |
| Rhode Island | 49,385 | 1,260 | 1,408,680 | 3.5% | – | – |
| South Carolina | 40,963 | 838 | 504,948 | 8.1% | 23,931 | 12.9% |
| South Dakota | 80,069 | 960 | 359,861 | 22.2% | 2,035 | 2.7% |
| Tennessee | 51,599 | 662 | 639,014 | 8.1% | 69,473 | 10.2% |
| Texas | 42,463 | 751 | 397,751 | 10.7% | 18,808 | 17.7% |
| Utah | 57,363 | 260 | 615,503 | 9.3% | 1,609 | 5.3% |
| Virginia | 26,817 | 472 | 433,495 | 6.2% | 5,675 | 9.5% |
| Washington | 21,159 | 357 | 380,093 | 5.6% | 2,844 | 3.8% |
| West Virginia | 24,652 | 397 | 602,108 | 4.1% | 1,537 | 4.2% |
| Wisconsin | 64,327 | 556 | 429,329 | 15.0% | 2,638 | 5.8% |
| Wyoming | 53,150 | 371 | 624,903 | 8.5% | 1,258 | 5.6% |

Data as of November 25, 2020 with 39,140 cases, 806 deaths, and 559,340 tests/1M in the United States for an overall 7.0% positive rate.
\*Testing as reported on the following websites: https://www.cdc.gov/covid-data-tracker/index.html#cases and www.worldometersinfo/coronavirus.
†Eurofins data based on PCR testing of 1,179,912 subjects in 47 states (39 of which with >100 cases are reported above) had a mean 9.3% positive rate as of November 1<sup>st</sup>, 2020. .
‡Pearson correlation between total reported state positive rates and US Eurofins Laboratories positive rates was $r = 0.609$ ( $P < 0.001$ ). 1M, 1 million people.

We also carried out a paired NP swab and saliva sample study, where we collected samples from previously positive subjects (n=91, mean age 53 years, 53% female) using an approved human institutional review board protocol (Trinity Health of New England, Hartford, CT); informed written consent was obtained from each subject. These subjects had all been positive 2-4 weeks earlier for SARS-CoV-2 RNA based on NP swabs. The saliva was collected in viral transport media using kits obtained from Strategic Laboratory Partners (Nashville, TN).

To compare our own primary data with data from the general population, we first accessed data as of September 1, 2020 and then subsequently on November 25, 2020 from the Centers for Disease Control and Prevention (CDC) website (https://www.cdc.gov/covid-data-tracker/index.html#testing), as well as the website https://www.worldometers.info/coronavirus/?, which uses a large number of data bases. For worldwide country data, we accessed the website https://www.worldometers.info/coronavirus/? and the World Health Organization website https://www.covid19.who.int/ on September 1, 2020 and again on November 25, 2020. On the latter date, there were 60,559,841 cases (7,769/million) and 1,423,530 deaths (183/million) worldwide. We specifically examined data for all countries with populations >50 million, as well as other selected countries. This analysis included 47 countries: 13 in Asia and Oceania; 11 in Europe; 5 in North America; 7 in South America; 7 in the Middle East; and 4 in Africa (Table 2).

**TABLE 2.** SARS-CoV-2 cases, deaths, and testing per million in the population

| Country | Cases/1 M | Deaths/1M | Tests/1M | % Positive | Case Fatality Rate |
| --- | --- | --- | --- | --- | --- |
| ASIA AND OCEANIA |  |  |  |  |  |
| India | 6,687 | 98 | 97,329 | 6.9% | 1.47% |
| Philippines | 3,839 | 75 | 50,714 | 7.5% | 1.95% |
| Indonesia | 1,863 | 59 | 19,899 | 9.4% | 3.17% |
| Bangladesh | 2,747 | 39 | 16,306 | 16.8% | 1.42% |
| Pakistan | 1,720 | 35 | 23,801 | 7.2% | 2.03% |
| Australia | 1,087 | 35 | 383,755 | 0.3% | 3.22% |
| Myanmar | 1,532 | 33 | 19,730 | 7.8% | 2.15% |
| South Korea | 619 | 10 | 57,839 | 1.1% | 1.64% |
| Japan | 1,072 | 16 | 26,373 | 4.1% | 1.49% |
| New Zealand | 408 | 5 | 248,599 | 0.2% | 1.23% |
| China | 60 | 3 | 111,163 | 0.05% | 5.00% |
| Thailand | 54 | 0.9 | 13,995 | 0.4% | 1.67% |
| Vietnam | 14 | 0.4 | 13,712 | 0.1% | 2.86% |
| EUROPE |  |  |  |  |  |
| Belgium | 48,390 | 1,373 | 492,764 | 9.8% | 2.84% |
| Spain | 34,700 | 942 | 468,697 | 7.4% | 7.40% |
| Italy | 24,507 | 861 | 346,808 | 7.1% | 3.51% |
| United Kingdom | 22,887 | 831 | 615,308 | 3.7% | 3.60% |
| France | 33,217 | 775 | 307,643 | 10.8% | 2.22% |
| Sweden | 22,768 | 647 | 287,225 | 7.9% | 2.84% |
| Ukraine | 15,171 | 263 | 97,269 | 15.6% | 1.73% |
| Russia | 14,815 | 257 | 504,934 | 1.7% | 2.93% |
| Germany | 11,678 | 182 | 315,370 | 3.7% | 1.56% |
| Finland | 4,086 | 70 | 335,210 | 1.2% | 1.71% |
| Norway | 6,250 | 58 | 401,259 | 1.6% | 0.93% |
| NORTH AMERICA |  |  |  |  |  |
| United States | 39,140 | 806 | 559,340 | 7.0% | 2.06% |
| Mexico | 8,188 | 794 | 21,186 | 38.6% | 9.70% |
| Panama | 36,145 | 688 | 195,453 | 18.5% | 1.91% |
| Canada | 9,121 | 309 | 291,154 | 3.1% | 3.39% |
| Guatemala | 6,649 | 228 | 28,759 | 23.1% | 3.43% |
| SOUTH AMERICA |  |  |  |  |  |
| Peru | 28,727 | 1,076 | 148,649 | 19.3% | 3.75% |
| Argentina | 30,462 | 825 | 81,851 | 37.2% | 2.71% |
| Brazil | 28,747 | 798 | 102,738 | 28.0% | 2.78% |
| Chile | 28,365 | 789 | 267,891 | 10.6% | 2.78% |
| Ecuador | 10,504 | 747 | 35,289 | 29.8% | 7.11% |
| Bolivia | 12,089 | 744 | 28,423 | 42.5% | 6.15% |
| Columbia | 24,701 | 698 | 120,080 | 20.6% | 2.80% |
| MIDDLE EAST |  |  |  |  |  |
| Iran | 10,594 | 547 | 70,045 | 15.1% | 5.16% |
| Israel | 36,061 | 307 | 597,111 | 6.0% | 0.85% |
| Iraq | 13,362 | 298 | 82,492 | 16.2% | 2.23% |
| Saudi Arabia | 10,165 | 166 | 268,395 | 3.8% | 1.63% |
| Turkey | 5,522 | 152 | 209,366 | 2.6% | 2.75% |
| Qatar | 47,268 | 83 | 348,678 | 13.6% | 0.18% |
| Egypt | 1,103 | 64 | 9,700 | 11.4% | 5.80% |
| AFRICA |  |  |  |  |  |
| South Africa | 12,956 | 354 | 89,347 | 14.5% | 2.73% |
| Kenya | 1,462 | 26 | 15,771 | 9.3% | 1.78% |
| Ethiopia | 923 | 14 | 13,869 | 6.7% | 1.52% |
| Nigeria | 320 | 6 | 3,599 | 8.9% | 1.88% |

As of November 25, 2020, there were 60,559,841 cases (7,769/million) and 1,423,530 deaths (183/million) worldwide.
Data as reported on the following websites: https://www.who.org and www.worldometersinfo/coronavirus.
Countries are grouped by continents and ranked according to number of deaths/million.

### SARS-CoV-2 viral detection

Detection of SARS-CoV-2 RNA in NP, OP, nasal swabs or saliva was performed using polymerase chain reaction (PCR) methods. As previously described, the Viracor assay using a reverse transcriptase PCR and TaqMan chemistry, targeted two regions of the SARS-CoV-2 nucleo-capsid (N) gene.^8^ The Boston Heart Diagnostics SARS-CoV-2 RNA assay was very similar to the Viracor assay except that this assay used Thermo-Fisher TaqPath COVID-19 Combo kits (Waltham, MA) and targeted a region in the N gene, a region in the spike (S) glycoprotein gene, and a region in the ORF1 gene. For both assays, a positive value was defined as detection of SARS-COV-2 RNA at a cycle threshold ≤37 cycles. The Diatherix assay was based on nested, end-point PCR technology that allowed for SARS-CoV-2 RNA detection through target enrichment and amplification. All assays have received emergency use authorization from the Food and Drug Administration. The sensitivity and specificity of these assays compared in known positive and negative subjects was found to be >95%.

### Statistical analysis

All statistical analyses were performed using R software, version 3.6.0 (R Foundation, Vienna, Austria) for comparisons between rates, and the statistical significance of differences between groups was assess using the nonparametric Kruskal-Wallis method. Pearson correlation analysis was performed.

## RESULTS

### United States data

As shown in Table 1, of 1,179,912 subjects having NP, OP, or nasal swabs done between March 13 and November 1, 2020 at various sites throughout the United States, the mean positive rate was 9.3%. When we first tabulated these data on June 1, 2020, New York State had by far the highest percentage of positive subjects (43.5%); this rate decreased to 4.3%, when we included a total of 213,926 tests, mainly done for health screening and for nursing home residents and employees. In our total study population, 18.2% were ≥65 years of age, of whom 6.9% were positive compared to 10.5% in the <65-year age group. Therefore, in the population we tested, older people did not have a higher positivity rate; in fact, it was lower (*P*<0.0001), even though it has been well documented that elderly subjects have a significantly higher case fatality rate than younger subjects.

We noted a significant correlation for the percentage of positives (r=0.609, *P*<0.0001) between our data and the CDC state-wide data for the 39 states where we had >100 cases/state (Table 1). As of November 25, 2020, based on CDC data, the top ten states in the United States for cases/million (all >50,000/million) were, in order, 1) North Dakota, 2) South Dakota, 3) Iowa, 4) Wisconsin, 5) Nebraska, 6) Utah, 7) Montana, 8) Illinois, 9) Kansas, and 10) Minnesota. In contrast, the top ten states in terms of deaths/million were 1) New Jersey, 2) New York, 3) Massachusetts, 4) Connecticut, 5) Louisiana, 6) Mississippi, 7) Rhode Island, 8) North Dakota, 9) Illinois, and 10) South Dakota. In terms of testing per 1 million, the top ten states were 1) Rhode Island, 2) Massachusetts, 3) New York, 4) Connecticut, 5) Illinois, 6) Louisiana, 7) Maryland, 8) New Mexico, 9) Minnesota, and 10) 6) Mississippi, 7) Rhode Island, 8) North Dakota, 9) Illinois, and 10) New Jersey.

The ten lowest states in terms of cases/million were 1) Oregon, 2) Washington, 3) West Virginia, 4) Virginia, 5) Pennsylvania, 6) California, 7) Connecticut, 8) Massachusetts, 9) Maryland, and 10) Ohio (Table 1). Similarly, the ten lowest states in terms of deaths/million were 1) Oregon, 2) Utah, 3) Washington, 4) Wyoming, 5) Oklahoma, 6) Kentucky, 7) Virginia, 8) California, 9) North Carolina, and 10) Idaho. The ten lowest states in terms of tests/million were 1) Oregon, 2) Pennsylvania, 3) Kansas, 4) Colorado, 5) Alabama, 6) Arizona, 7) South Dakota, 8) Washington, 9) Idaho, and 10) Texas.

Figure 1 shows the relationship between death and case rates per million by state (Panel A), as well as the relationship between the testing rate and the case rate (Panel B), both on a linear scale. These data from the CDC are based on 39,140 cases/million, 806 deaths/million and 559,370 tests/million in the US population. As can be clearly seen, the northeastern states of New Jersey, New York, Massachusetts, Connecticut, and Rhode Island had very high death rates per case. Intermediate mortality per case were observed in the southern states and the midwestern states. The lowest mortality per case were seen in the western states, especially Oregon, Utah, Washington, and Wyoming. As of September 1, 2020, the overall correlation, using Pearson correlation coefficient analysis, between deaths/million and cases/million was 0.473 (*P*<0.0001); between tests/million and cases/million, it was 0.398 (*P*<0.0001).^9^ By November 25, 2020, the correlations had decreased significantly to 0.076 and -0.093, respectively (Figure 1). In our view, these changes relate to more testing and less population public health measures. The states where the governors introduced early and constant public health measures had the lowest case and death rates, and the converse was also true.

**FIGURE 1.**
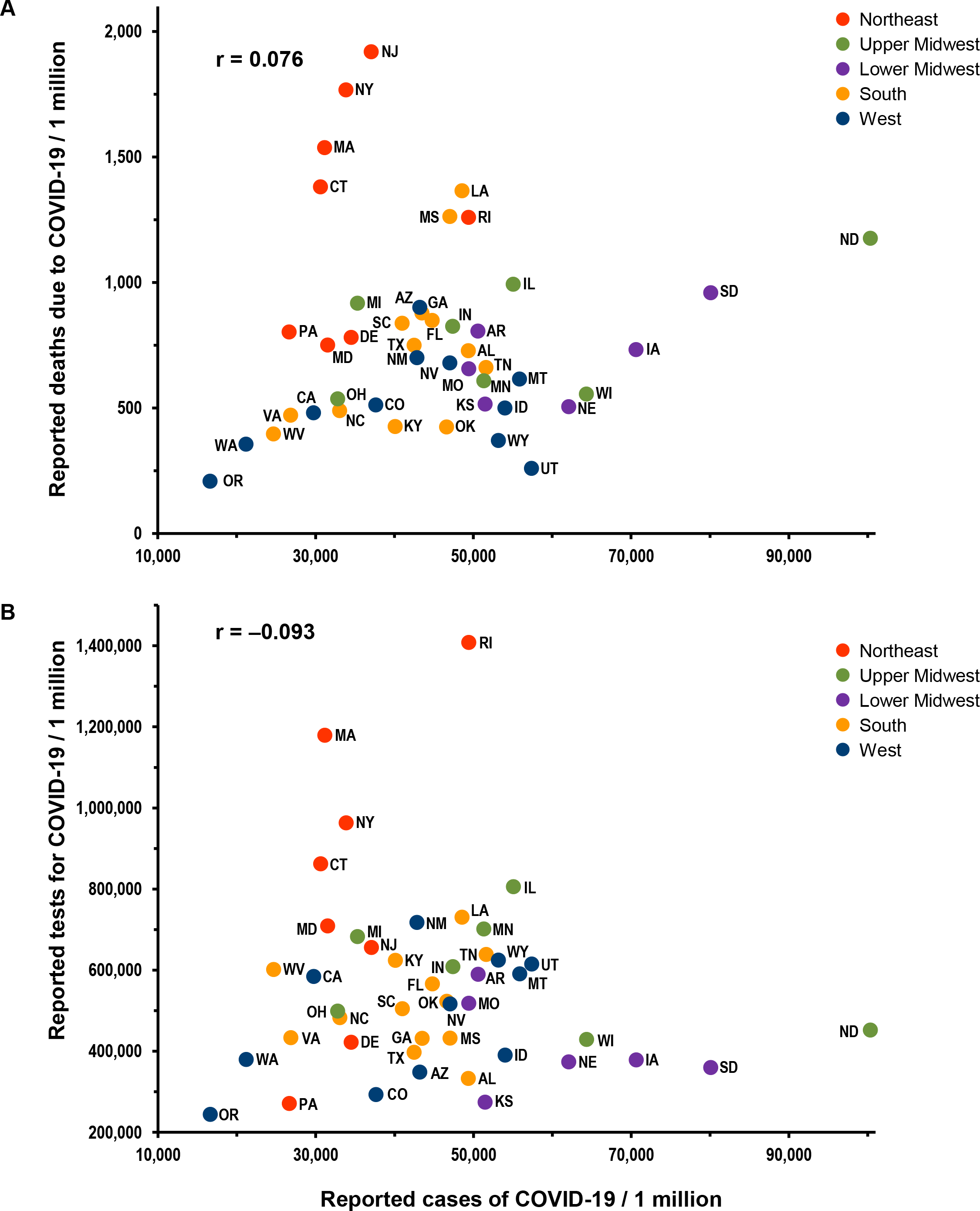
COVID-19 death and testing rates relative to case rates in the United States as of November 25, 2020. The reported cases of COVID-19 per 1 million people are presented by state versus the reported deaths due to COVID-19 (Panel A) and versus the reported tests for COVID-19 (Panel B). Red circles indicate states in the Northeast; green circles, states in the upper Midwest; purple circles, states in the lower Midwest; orange circles, states in the South; and blue circles, states in the West. Linear scales were used.

### Paired NP swab and saliva data

In the paired analysis examining positivity rates for NP swabs and saliva samples in 91 previously positive subjects, we noted that 58.6% of subjects were still positive based on NP swabs, but only 21.5% were positive based on saliva collection. These differences were statistically significant *P*<0.01). We also documented that NP swabs could remain positive for 6 weeks or longer in this analysis in some cases.

### Worldwide data

Data on the relationship between deaths/million and cases/million and between tests/million and cases/million are shown in Figure 2. These data are plotted on a log scale because of the marked variability between countries. As of September 1, 2020, the correlation between cases/million and deaths/million (0.488, *P*<0.0001) was similar to what we observed for the US states, as were the correlation between cases/million and tests/million (0.395; *P*<0.0001). However, as of November 25, 2020, the correlations worldwide had moved in the opposite direction. The correlations between cases/million and deaths/million had increased to 0.763 (*P*<0.0001) and those between cases/million and tests/million had increased to 0.600(*P*<0.0001).

**FIGURE 2.**
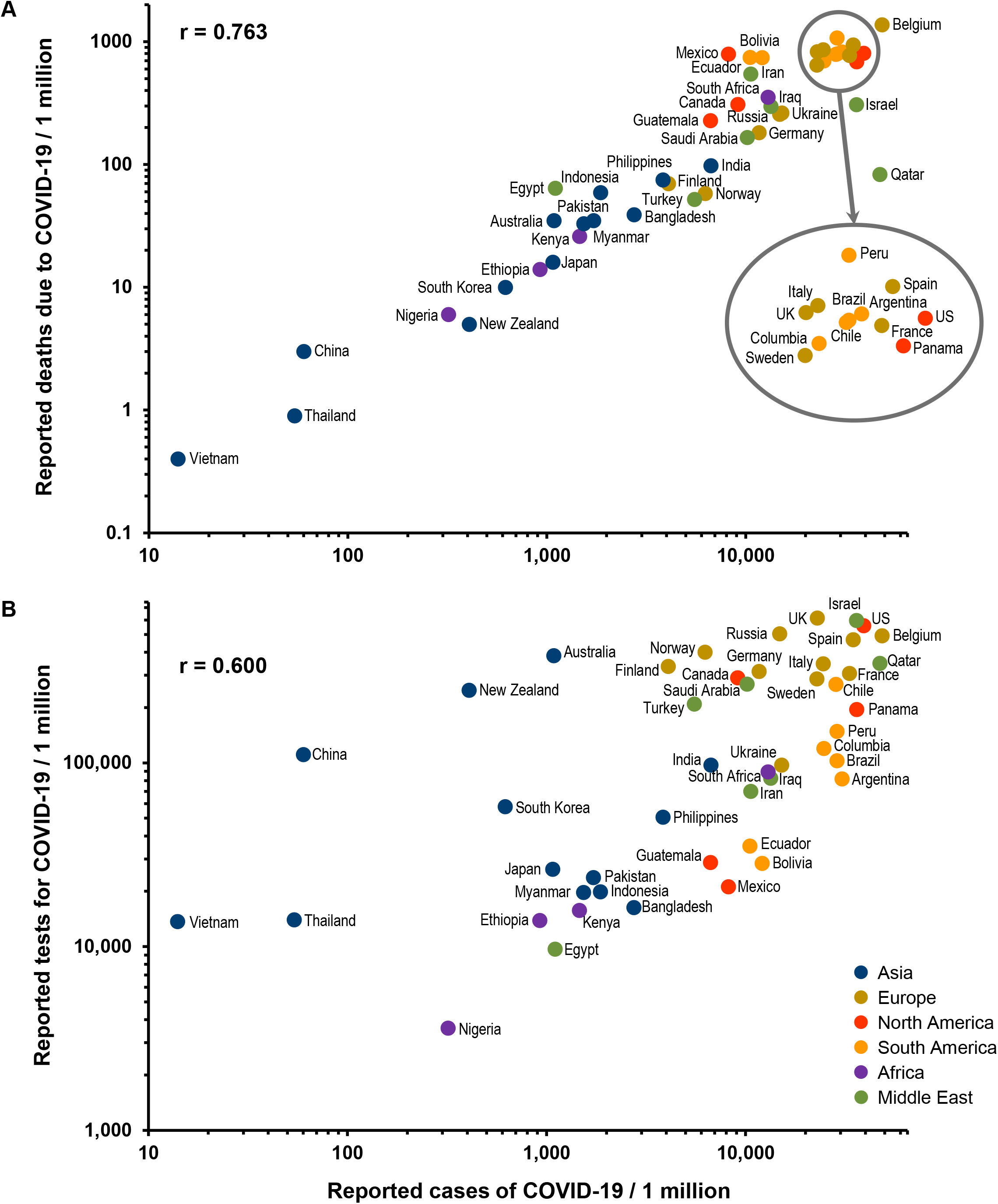
COVID-19 fatality and testing rates relative to case rates world-wide as of November 25, 2020. The reported cases of COVID-19 per 1 million people are presented by country versus the reported deaths due to COVID-19 (Panel A) and versus the confirmed tests for COVID-19 (Panel B). Blue circles represent Asia; gold circles, Europe; red circles, North America; orange circles, South America, purple circles, Africa; and green circles, the Middle East. Log scales were used. UK, United Kingdom, US, United States.

As of November 25, 2020, as shown in Table 2, the top ten countries for cases/million in order were: 1) Belgium, 2) Qatar, 3) United States, 4) Panama, 5) Israel, 6) Spain, 7) France, 8) Argentina, 9) Brazil, and 10) Peru. The top ten countries in terms of deaths/million in order were: 1) Belgium, 2) Peru, Spain, 4) Italy, 5) United Kingdom, 6) Argentina, 7) United States, 8) Brazil, 9) Mexico, and 10) Chile. In terms of testing per 1 million, the top countries in order were: 1) United Kingdom, 2) Israel, 3) United States, 4) Russia, 5) Belgium, 6) Spain, 7) Norway, 8) Italy, 9) Finland, and 10) Germany.

The ten countries with the lowest cases/million in order were: 1) Vietnam, 2) Thailand, 3) China, Nigeria, 5) New Zealand, 6) South Korea, 7) Ethiopia, 8) Japan, 9) Australia, and 10) Myanmar. Similarly, the ten countries with the lowest deaths/million in order were: 1) Vietnam, 2) Thailand, 3) China, 4) New Zealand, 5) Nigeria, 6) South Korea, 7) Ethiopia, 8) Japan, 9) Kenya, and 10) Myanmar. The ten countries with the lowest tests/million in order were: 1) Nigeria, 2) Egypt, 3) Ethiopia, 4) Vietnam, 5) Thailand, 6) Bangladesh, 7) Myanmar, 8) Indonesia, 9) Mexico, and 10) Japan.

## DISCUSSION

The rapid spread of SARS-CoV-2 viral infection has been in large part due to its very contagious nature and the fact that many infected people are asymptomatic. Although the pandemic started in China, its spread there, as well as in other countries in Asia, has been very well controlled; and the case and death rates per million in these countries have been very low. One cannot attribute this excellent infection control to testing, but rather to outstanding public health measures (use of face masks, isolation, contact tracing, and social distancing). Even in countries such as India, Bangladesh, the Philippines, and Australia with >1,000 cases/million, death rates have been low. In contrast, subjects in Europe, North America, South America, and parts of the Middle East have fared far worse with much higher case and death rates, despite a large amount of testing. In the midwestern and southern states of the United States case rates have been very high, while death rates have been the highest in the world in the northeastern states, despite a lot of testing. This latter finding may well have been due to the high infection rate early on in the United States pandemic in long-term care facilities in the northeast. Recently, there has been a large decrease in the correlation between cases and testing in the United States, indicating a lot of testing, but lack of public health measures, especially in younger people participating in demonstrations and political rallies.^9^

Rates in the United States and Brazil were comparable in cases and deaths, due in our view to very limited public health measures and lack of central government leadership in both countries. In contrast, Japan, South Korea, China, and Thailand had much lower rates, presumably due to significantly better public health measures. Many European countries had case rates lower than the United States, but comparable death rates. These differences could relate to the age of subjects becoming infected. The importance of public health measures may best be exemplified by comparing Sweden, which did not introduce such measures, to its neighbors Norway and Finland, which did introduce such measures. Sweden had case and death rates that were much higher than rates observed in Norway and Finland. These marked differences occurred despite the fact that these countries had fairly comparable testing rates.

There may be other potential causes of the large variability in case and death rates between countries. One such possibility is mutations in the virus. The D614G amino acid substitution in the S glycoprotein as well as other variants encoded by the SARS-CoV-2 S gene has been reported to result in forms of the virus that may be more infective and virulent than the original Wuhan strain.^10-16^ The D614G variant has been found in over 90% of United States strains. Another possibility is human genetic variation. Genome-wide association studies have identified a 3p21.31 DNA locus as being associated with a significant 1.77-fold increased risk for respiratory failure in hospitalized COVID-19 patients.^17^ This genetic variant, apparently inherited from Neanderthals, is not present in subjects indigenous to China, Japan, or Sub-Saharan Africa, but has a frequency of ∼5−8% in North America and western Europe and ∼20% in India.^18^ Therefore, both viral and human genetic variation may play a role in country differences with regard to SARS-CoV-2 cases and mortality rates. Another potential reason for mortality differences is the age of infected people, since it is well known the elderly have much higher SARS-CoV-2 mortality rates than the young. However, it is most likely that the efficiency of public health measures accounts for most of the variability in case and death rates per million in the population.^19,20^

Recently, there has been a significant increase in SARS-CoV-2 RNA testing in the United States in an effort to control the pandemic in the absence of vaccines. There have also been efforts to find easier ways to carry out such testing. Data from Yale New Haven Medical Center indicated that self-collected saliva samples yielded similar or better results than did NP swabs in terms of detecting SARS-CoV-2 positive hospitalized patients based on 44 paired samples.^3^ However, our data in previously positive outpatients indicated that saliva analysis only identified about half as many positive cases as compared to NP swab analysis. Saliva testing is being widely used.

## CONCLUSIONS

Our data indicate that: 1) outpatient saliva testing is not as sensitive as NP testing; 2) the marked variability in case and death rates between states and countries is mainly due to difference in public health measures; 3) variations in SARS-CoV-2 viral genetics, human genetics, and age of populations getting infected may also play a role in rate differences; and 4) rate differences are least likely to be due differences in testing rates. Our overall data strongly support the benefits of public health measures in preventing spread of SARS-CoV-2 infection. In our view, the major reason for the very high case and mortality rates in the United States, as well as some other countries, has been the consistent lack of such measures, due to failures of central government and public agency leadership.

## Data Availability

The raw data from this study are available upon request to the corresponding author.
Information on COVID-19 data for the United States was obtained from the Centers for Disease Control and Prevention (CDC) website on September 1, 2020, https://www.cdc.gov/covid-data-tracker/index.html#testing, as well as from https://www.worldometers.info/coronavirus/? and https://www.covid19.who.int/.

https://www.worldometers.info/coronavirus/?

https://www.covid19.who.int/

## Abbreviations

CDC: Centers for Disease Control and Prevention
COVID-19: coronavirus disease-2019
EUA: emergency use authorization
FDA: Food and Drug Administration
N gene: nucleocapsid gene
NP: naso-pharyngeal
OP: oro-pharyngeal
PCR: polymerase chain reaction
RT-PCR: reverse transcriptase polymerase chain reaction
SARS-CoV-2: severe acute respiratory syndrome coronavirus-2
S gene: spike glycoprotein gene

## Acknowledgements

We thank the medical personnel and technical staff at each institution for their effort and commitment to SARS-CoV-2 testing. We also thank Shannon Foster of Viracor-Eurofins Clinical Diagnostics and Scientific Network for compiling all RNA data. All information was anonymized prior to data analysis. All relevant ethical guidelines were followed in carrying out this research.

## Research Support and Disclosures

Support for this research was received from Boston Heart Diagnostics, Viracor-Eurofins Clinical Diagnostics and Scientific Network, Diatherix, and Trinity Health of New England. No relevant conflicts of interest were identified, except that many authors are either full-time or part-time employees of reference laboratories that provide testing for healthcare providers [Boston Heart Diagnostics (EJS, ASG, MRD), Diatherix (JW), and Viracor-Eurofins Clinical Diagnostics (SBK)]. Boston Heart Diagnostics, Diatherix, and Viracor-Eurofins Clinical Diagnostics are part of the global network of Eurofins laboratories and the Eurofins Scientific Network with headquarters in Brussels, Belgium. No payments were received from any third party to influence the results of the research.

## Notes

### Competing Interest Statement

The authors have declared no competing interest.

### Clinical Trial

not a clinical trial

### Author Declarations

All subject information was anonymized prior to data analysis, and all relevant ethical guidelines were followed. We have followed the STROBE Guidelines for Observation Studies. IRB oversight for patient data extracted from medical records without name or identification number and analyzed as anonymized data was provided by Advarra Institutional Review Board (Columbia, MD). As stated in the cover letter and in the manuscript, the determination of the Advisory Review dated 25. September 2020 was "had the request for exempt determination been submitted prior to initiation of research activities, the research would have met the criteria for exemption from IRB review under 45 CFR 46.104(d)(4)." The study population included a subset of 91 previously diagnosed COVID-19 subjects who were being evaluated as potential participants in an approved human IRB protocol at Trinity Health of New England (Hartford, CT).

